# COVID-19 Vaccine Hesitancy in Ghana: The Roles of Political Allegiance, Misinformation Beliefs, and Sociodemographic Factors

**DOI:** 10.1101/2022.03.16.22272463

**Authors:** Ken Brackstone, Kirchuffs Atengble, Michael G Head, Laud A Boateng

## Abstract

The vast majority of people in the world who are unvaccinated against COVID-19 reside in LMIC countries in sub-Saharan Africa. This includes Ghana, where only 15.9% of the country are fully vaccinated as of April 2022. A key factor negatively impacting vaccination campaigns is vaccine hesitancy, defined as the delay in the acceptance, or blunt refusal, of vaccines. Four online cross-sectional surveys of Ghanaian citizens were conducted in August 2020 (*N* = 3048), March 2021 (*N* = 1558), June 2021 (*N* = 1295), and February 2022 (*N* = 424) to observe temporal trends of vaccine hesitancy in Ghana, and to examine key groups associated with hesitancy. Overall hesitancy decreased from 36.8% (95% CI: 35.1%-38.5%) in August 2020 to 17.2% (95% CI: 15.3%-19.1%) in March 2021. However, hesitancy increased to 23.8% (95% CI: 21.5%-26.1%) in June 2021, and then again to 52.2% (95% CI: 47.4%-57.0%) in February 2022. Key reasons included not having enough vaccine-related information (50.6%) and concerns over vaccine safety (32.0%). Hesitant groups included Christians, urban dwellers, opposition political party voters, people with more years of education, females, people who received COVID-19 information from internet sources, and people who expressed uncertainty about COVID-19 misinformation beliefs.

## 1. Introduction

While high-income settings have achieved relatively high coverage with their COVID-19 vaccination campaigns, 35% of the world’s population are still yet to receive a single dose of any COVID-19 vaccine as of April 2022 (Ritchie et al., 2022). The vast majority of unvaccinated people reside in lower-income settings in sub-Saharan Africa (SSA). This includes Ghana, a country in West Africa with an estimated 30.8m population. Ghana has reported over 161,000 cases and 1445 deaths, and currently only 19.7% of the country is considered fully vaccinated (Ritchie et al., 2022). With the emergence of the highly-transmissible Omicron variant (Wolter et al., 2021), large-scale vaccination coverage is fundamental to the national and global pandemic response.

Three common factors that impact the success of vaccination campaigns include inequity around supply and demand issues, social mobilization and logistical issues (challenges often related to underfunded health programmes), and vaccine confidence once doses arrive in communities. Vaccine hesitancy is defined by the World Health Organization (WHO) as the delay in the acceptance, or blunt refusal of, vaccines. Hesitancy was described by the WHO as one of the top 10 threats to global health in 2019, and has been identified as a growing trend in SSA more generally (Marti et al., 2017). Developing a deeper understanding of the factors associated with vaccine hesitancy is crucial toward informing locally-tailored health promotion strategies. Hesitancy in SSA has previously been associated with mistrust, particularly of government messaging, and the presence of misinformation. Examples include a boycott of the polio vaccine in Northern Nigeria in 2003-2004 (Jegede, 2007), as well as more recent surveys administered in SSA countries such as Malawi, Mali, and Nigeria, which found that dissatisfaction with the government’s response to the COVID-19 pandemic predicted hesitancy (Kanyanda et al., 2021). Vaccine misinformation is also amplified on social media (Dotto & Cubbon, 2021), especially from specific faith institutions, individuals, and other organized groups that have become deeply entrenched in online platforms such as Facebook (Puri & Coomes, 2020).

A handful of research has described COVID-19-related vaccine hesitancy in SSA countries (Kanyanda et al., 2021; Olapegba et al., 2020), but overall, there is still a limited evidence base around COVID-19-related vaccine hesitancy in sub-Saharan Africa and specifically within Ghana. This study presents estimates of vaccine hesitancy in Ghana from four nationally-representative online samples of Ghanaian residents that took place between August 2020 and June 2021. The study describes the groups most associated with vaccine hesitancy using demographic and socioeconomic variables, including political allegiance and misinformation beliefs.

## 2. Materials and Methods

### 2.1. Study design

Four cross-sectional surveys were administered. Survey 1 was conducted in August 2020, three months after the first case of COVID-19 was reported in Ghana and prior to any vaccines being globally approved and available. Survey 2 was conducted in March 2021 shortly after the first batch of Oxford AstraZeneca (AZ) vaccines arrived in Ghana through the COVAX initiative and the beginning of vaccine rollout. Survey 3 was conducted in June 2021, three months after vaccine rollout begun in Ghana. Finally, Survey 4 – intended as a short survey to detect up-to-date hesitancy trends in Ghana – was conducted in February 2022. Participants completed a self-administered online survey using Qualtrics XM. Each survey was available online for approximately 4 weeks. Dissemination was conducted using a snowball effect of word-of-mouth (Whatsapp, LinkedIn, email, direct messaging), and advertising via Facebook Ads Manager. This technique allowed us to direct the survey toward individuals whose Facebook profile was registered as them being aged 18 and over and residing anywhere in Ghana. Associated study information appeared on individuals’ Facebook timelines along with the survey link. In each survey, participants were offered the opportunity to enter a prize draw to win 1 of 25 money vouchers worth 100 Ghana Cedis (approximately 12 GBP) upon full completion of the survey. Qualtrics allows checks to ensure that consented participants could only take the survey once from the same IP address. Sample size calculations suggested 1067 participants in each survey, providing approximately 3% margin of error at 95% confidence. The surveys were conducted in English.

Informed consent was required to participate in the study after reading the study information statement online. Participants read the study aims and design displayed online, and then checked a tick box to confirm consent. The surveys received ethical approvals from University of Southampton Ethics Committee (Institutional Review Board ID: 57267) and conformed to local ethical standards applied according to the Ghana Health Service (GHS), Ghana.

### 2.2. Measures

#### Vaccine hesitancy

In Survey 1, participants were asked: “When the COVID-19 vaccine becomes available to you, would you like to get vaccinated?” (yes, no, I don’t know). In Surveys 2-4, participants initially indicated whether they had previously received any doses of the COVID-19 vaccine. Among participants who indicated that they had not received any doses, vaccine hesitancy was then assessed using two distinct measurements. First, participants were asked: “When the COVID-19 vaccine becomes available to you, would you like to get vaccinated?” (yes, no, I don’t know). Participants who indicated disagreement or indecision about receiving the vaccine subsequently specified reasons for their hesitancy. A list of nine reasons was consequently presented□ (e.g., “[COVID-19] is not serious enough to need a vaccine”). Second, participants indicated the extent of their agreement to the question: “If a vaccine for COVID-19 were available to me, I would get it.” (1 = *strongly disagree*, 5 = *strongly agree*; *M* = 4.02, *SD* = 1.33).

Participants indicated whether they knew anybody personally who had received the COVID-19 vaccine (yes, no).

#### Misinformation about COVID-19

Participants indicated whether they believed in seven COVID-19-related misinformation beliefs recorded to be circulating in sub-Saharan Africa by selecting ‘yes’ if they agreed with the belief, ‘unsure’ if they were uncertain about the belief, or ‘no’ if they did not agree with the belief (e.g., “To the best of your knowledge… [COVID-19] is designed to reduce or control the population”; Olapegba et al., 2020).

#### Sources of COVID-19 vaccine-related information

Participants selected (yes or no) where they typically retrieved COVID-19 vaccine-related information from a list of eight sources (Olapegba et al., 2020). These included common social media platforms (Facebook, Whatsapp, Twitter), more traditional news sources (TV/radio, Ghana Health Service [GHS]), government officials, the internet (e.g., news websites), and family members and friends.

#### Political allegiance

Participants selected the political party that they voted for in the Ghanaian election of December 2020 (New Patriotic Party [NPP; elected], National Democratic Congress [NDC; unelected], other, none/I didn’t vote). Participants indicated their trust in the vaccine (“I would trust the safety of the COVID-19 vaccine when it becomes available to me,” 1 = *strongly disagree*, 5 = *strongly agree*; *M* = 3.96, *SD* = 1.29). Then, using a 2-item measure, confidence in the current government was assessed (e.g., “I have trust in the Ghanaian government’s response to the COVID-19 pandemic;” 1 = *strongly disagree*, 5 = *strongly agree*), which was averaged to form a governmental response index (*r* = .80, *M* = 3.62, *SD* = 1.32).

#### Demographic variables

Finally, participants indicated their age, gender, marital status, and religion. Socioeconomic variables included employment status, education, and community type (urban, rural).

### 2.3. Data analysis

The data captured in Qualtrics were examined for errors, cleaned, and exported into IBM SPSS Statistics 28 for further analysis. Descriptive statistics summarized respondents’ socio-demographics. Inferential statistics were conducted in three phases. First, temporal trends in hesitancy and population prevalence were compared between each survey. For consistency between surveys, hesitancy was coded by dichotomising participants’ responses (no, I don’t know) to the question: “When the COVID-19 vaccine becomes available to you, would you like to get vaccinated?” Chi-Square χ^2^ tests were conducted to assess for categorical differences in hesitancy rates between Surveys 1-2, 2-3, and 3-4. Descriptive analyses were also conducted to summarize misinformation beliefs and self-reported sources of vaccine-related information.

In Survey 3, bivariate logistic regressions assessed relationships between individual predictors and vaccine hesitancy (S1). A combined logistic regression model was then administered containing all predictors in a single model, providing the strictest test of potential associations with vaccine hesitancy. To account for the level of variance in participants’ responses, vaccine hesitancy was coded by dichotomising participants’ responses (strongly disagree, somewhat disagree, or undecided) to the statement: “If a vaccine for COVID-19 were available to me, I would get it.” Vaccine hesitancy and its associated predictors were rescaled to 0 or 1 in our statistical analyses, which allowed for direct comparison of effect sizes.

Finally, a one-way ANOVA and associated post-hoc tests were conducted to compare differences in political groups’ ratings of trust in the COVID-19 vaccine.

## 3. Results

### 3.1. Participant characteristics

Table 1 shows descriptive statistics of participants from Survey 1 (N = 3048), Survey 2 (N = 1558), Survey 3 (N = 1295), and Survey 4 (424). Among the largest ethnic groups in this sample were Akan (46.3%) and Ewe (16.4%). The majority of participants lived in Greater Accra (28.5%) and Ashanti (17.9%) regions. Further, 62.3% had completed higher education, and 37.3% had completed senior secondary education or lower. More than 60% of participants reported being single (62.0%) vs. married or in a relationship (37.0%), 58.7% reported living in an urban community (vs. rural; 39.0%), and 45.7% reported being unemployed (vs. 52.0% employed to some degree), while 82.5% reported being Christian (vs. 17.5% Muslim). Finally, 71.7% reported knowing someone personally who had received the vaccine.

**Table 1.**
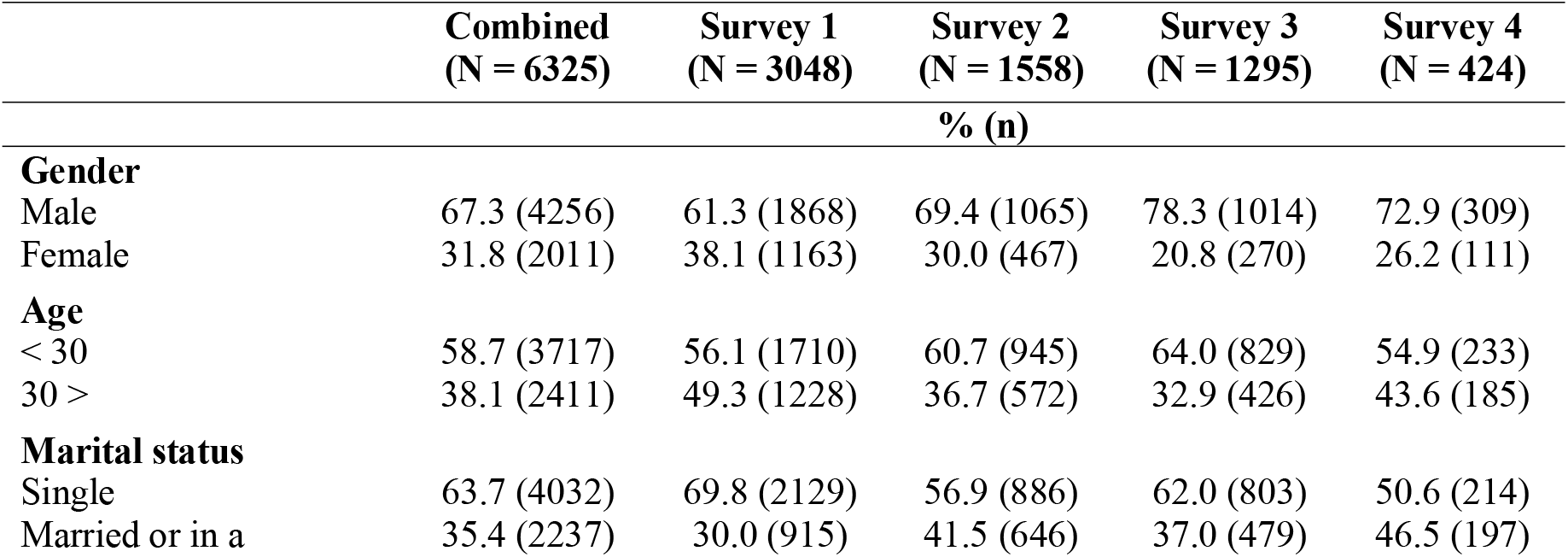
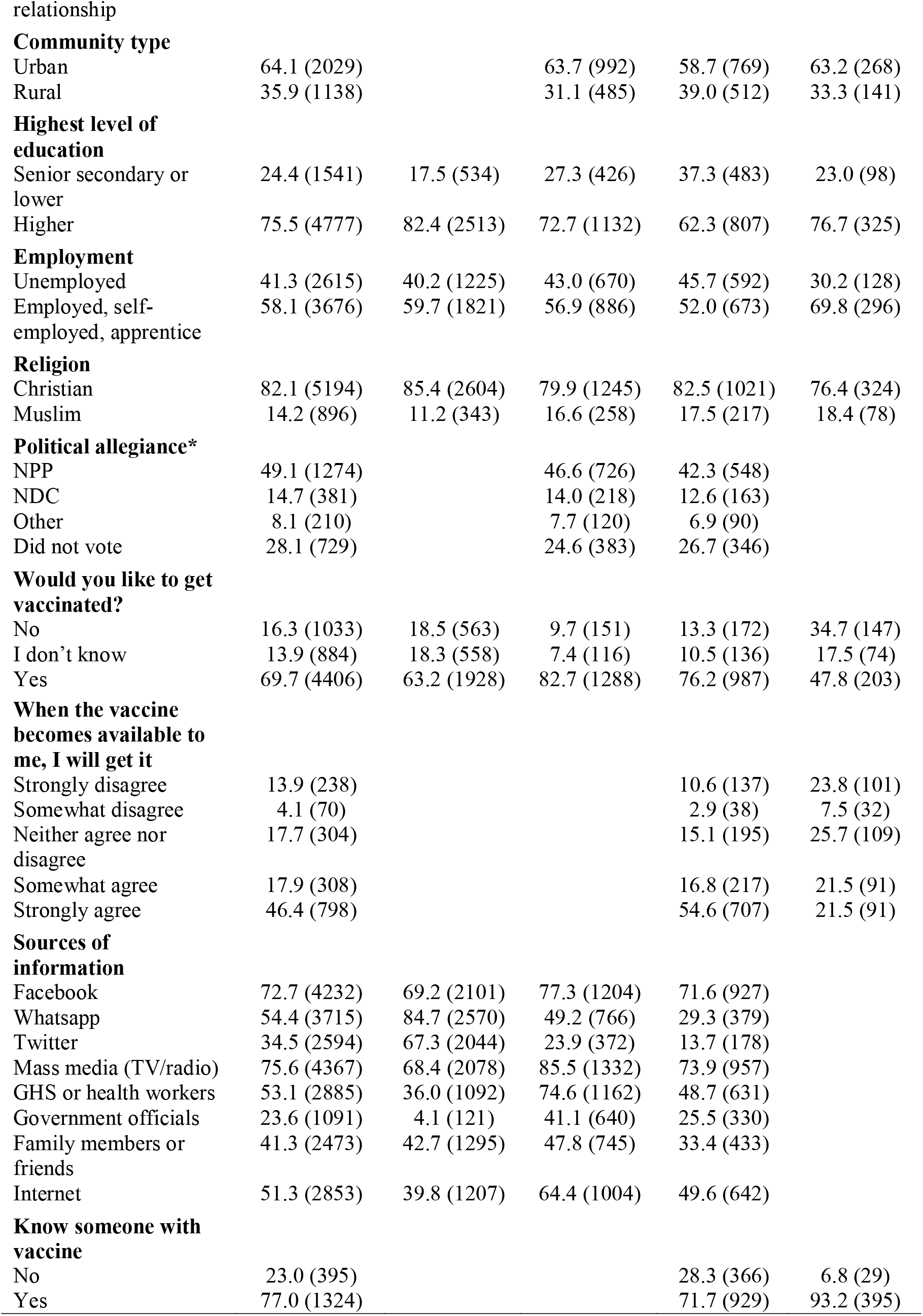
Descriptive statistics of participants from 4 surveys of data collection. Empty cells indicate that these variables were not collected. *Note:* Percentages may not equal 100% due to incomplete questions.

### 3.2. Hesitancy trends across time

A Pearson’s Chi-Squared test revealed a significant association (Figure 1) between time and vaccine hesitancy (χ2 (1) = 182.687 *p* < .0001), in which overall hesitancy decreased from 36.8% (CI: 35.1%-38.5%) in Survey 1 (August 2020) to 17.2% (95% CI: 15.3%-19.1%) in Survey 2 (March 2021). Hesitancy increased (χ2 (1) = 19.188, *p* < .0001) from 17.2% in Survey 2 to 23.8% (95% CI: 21.5%-26.1%) in Survey 3 (June 2021). Finally, hesitancy increased again (χ2 (1) = 120.413, *p* < .0001) from 23.8% in Survey 3 to 52.2% (95% CI: 47.4%-57.0%) in Survey 4 (February 2022).

**Figure 1.**
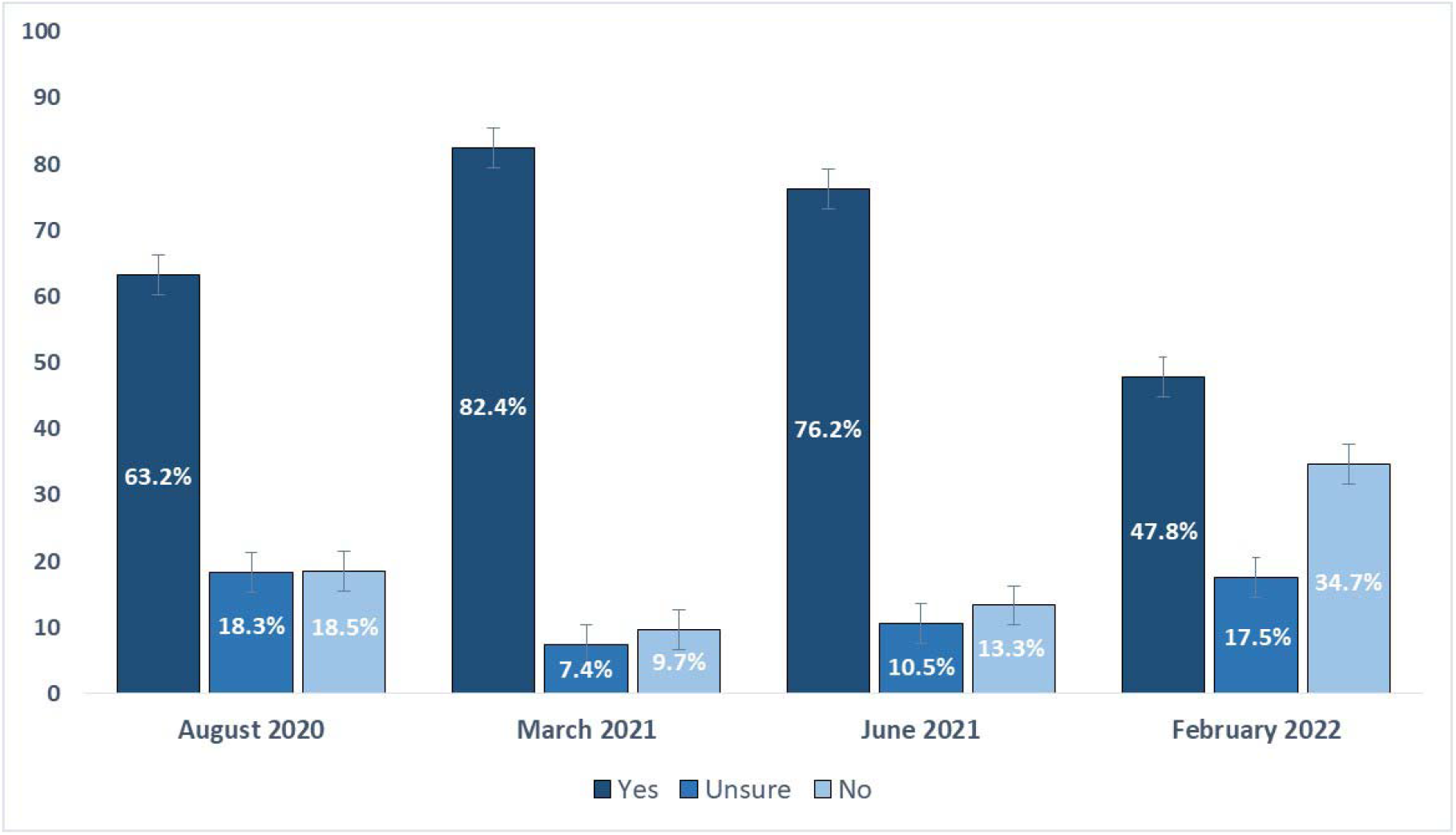
Breakdown of yes, no, and unsure responses across four surveys in August 2020, March 2021, June 2021, and February 2022.

### 3.3. Predictors of vaccine hesitancy

The following analyses derive from Survey 3 (June 2021), which measured more useful predictors to include in our regression model. Among participants who indicated ‘no’ and ‘I don’t know’ that they would take the vaccine when available (308/1295; 23.8%), the most common reasons were: not having enough information about vaccine (156/308; 50.6%), believing that the vaccine would be unsafe or dangerous (99/308; 32.1%), not trusting the government or service departments (66/308; 21.4%), and believing that they would experience side effects and get sick from the vaccine (58/308; 18.8%; Table 2)

**Table 2.** Reasons for refusing the vaccine in March 2021, June 2021, and February 2022.

|  | Survey 2 (N =<br>267/1558) | Survey 3 (N =<br>308/1295) | Survey 4 (N =<br>221/424) |
| --- | --- | --- | --- |
|  | n (%) |  |  |
| **The vaccine will be dangerous** |  |  |  |
| Yes | 24.0 (64) | 32.1 (99) | 47.5 (105) |
| No | 76.0 (203) | 67.9 (209) | 52.5 (116) |
| **I will experience side effects and get sick** |  |  |  |
| Yes | 30.1 (82) | 18.8 (58) | 33.5 (74) |
| No | 69.3 (185) | 81.2 (250) | 66.5 (147) |
| **COVID-19 is not severe enough to need a vaccine** |  |  |  |
| Yes | 15 (5.6) | 14.0 (43) | 27.6 (61) |
| No | 252 (94.4) | 86.0 (265) | 72.4 (160) |
| **I don't trust the government department** |  |  |  |
| Yes | 22.8 (61) | 21.4 (66) | 16.3 (69) |
| No | 77.2 (206) | 78.6 (242) | 152 (83.7) |
| **I will be allergic to the vaccine** |  |  |  |
| Yes | 9.7 (26) | 7.1 (22) | 11.8 (26) |
| No | 90.3 (241) | 92.9 (286) | 88.2 (195) |
| **It is too far to travel to the vaccination centre** |  |  |  |
| Yes | 1.9 (5) | 6.8 (21) | 11.8 (26) |
| No | 98.1 (262) | 93.2 (287) | 88.2 (195) |
| **The vaccine will not work** |  |  |  |
| Yes | 9.7 (25) | 10.4 (32) | 23.5 (52) |
| No | 90.6 (242) | 89.6 (276) | 76.5 (169) |
| **I don't have enough information about the vaccine** |  |  |  |
| Yes |  | 50.6 (156) | 52.0 (115) |
| No |  | 49.4 (152) | 48.0 (106) |
| **I don't need it** |  |  |  |
| Yes |  | 13.0 (40) | 19.5 (43) |
| No |  | 87.0 (268) | 80.5 (178) |

In all, 44.2% (573/1295) of participants indicated agreement with at least one of the seven widely circulated misinformation beliefs (*M* = 1.38, *SD* = 1.97; Table 3). The most common misinformation beliefs included: “… is a biological weapon caused by the Chinese government” (316/1295; 24.4%), “… is a result of 5G technology being installed in Ghana” (310/1295; 23.9%), and “… is a biological weapon designed by the U.S. government” (297/1295; 22.9%).

**Table 3.**
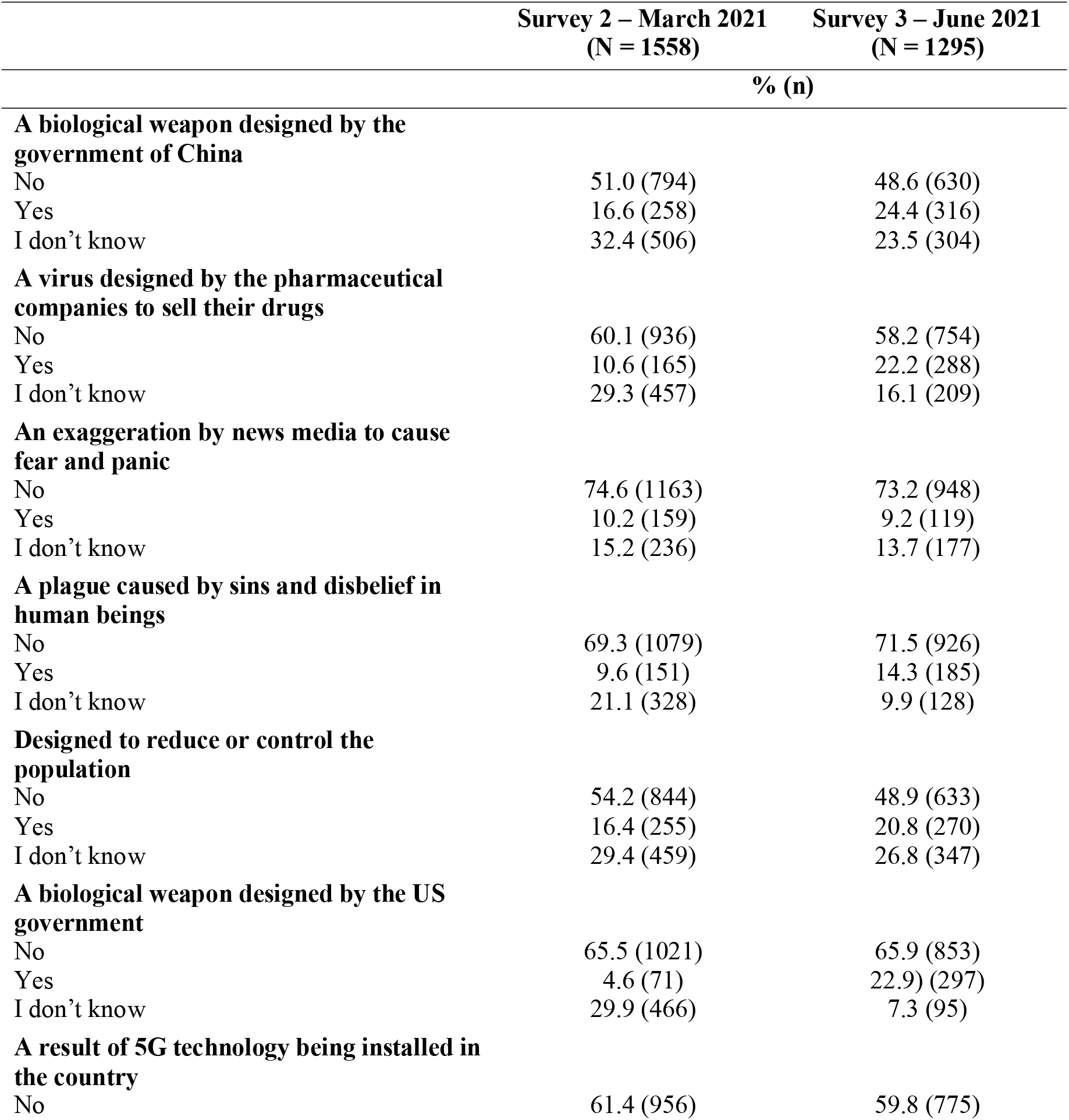
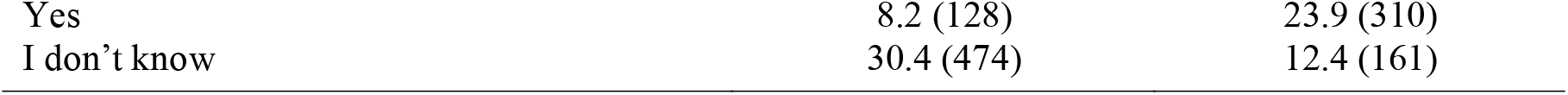
Breakdown of COVID-19 misinformation beliefs in Surveys 2 and 3. *Note:* Percentages may not equal 100% due to incomplete questions.

|  | Survey 2 – March 2021<br>(N = 1558) | Survey 3 – June 2021<br>(N = 1295) |
| --- | --- | --- |
|  | % (n) |  |
| **A biological weapon designed by the government of China** |  |  |
| No | 51.0 (794) | 48.6 (630) |
| Yes | 16.6 (258) | 24.4 (316) |
| I don't know | 32.4 (506) | 23.5 (304) |
| **A virus designed by the pharmaceutical companies to sell their drugs** |  |  |
| No | 60.1 (936) | 58.2 (754) |
| Yes | 10.6 (165) | 22.2 (288) |
| I don't know | 29.3 (457) | 16.1 (209) |
| **An exaggeration by news media to cause fear and panic** |  |  |
| No | 74.6 (1163) | 73.2 (948) |
| Yes | 10.2 (159) | 9.2 (119) |
| I don't know | 15.2 (236) | 13.7 (177) |
| **A plague caused by sins and disbelief in human beings** |  |  |
| No | 69.3 (1079) | 71.5 (926) |
| Yes | 9.6 (151) | 14.3 (185) |
| I don't know | 21.1 (328) | 9.9 (128) |
| **Designed to reduce or control the population** |  |  |
| No | 54.2 (844) | 48.9 (633) |
| Yes | 16.4 (255) | 20.8 (270) |
| I don't know | 29.4 (459) | 26.8 (347) |
| **A biological weapon designed by the US government** |  |  |
| No | 65.5 (1021) | 65.9 (853) |
| Yes | 4.6 (71) | 22.9 (297) |
| I don't know | 29.9 (466) | 7.3 (95) |
| **A result of 5G technology being installed in the country** |  |  |
| No | 61.4 (956) | 59.8 (775) |
| Yes | 8.2 (128) | 23.9 (310) |
| I don't know | 30.4 (474) | 12.4 (161) |

Next, 46.4% (601/1295) of participants indicated uncertainty about at least one COVID-19-related misinformation belief; *M* = 1.10, *SD* = 1.55). The most common misinformation beliefs in which participants indicated uncertainty included: “… was designed to reduce or control the population” (347/1295; 26.8%), “… is a biological weapon caused by the Chinese government” (304/1295; 23.5%), and “… is a virus designed by the pharmaceutical industry to sell their drugs” (209/1295; 16.1%).

The most commonly accessed sources of COVID-19 vaccine-related information were mass media (e.g., newspapers, radio, TV; 73.9% [957/1295]), Facebook (77.3% [927/1295]), the Internet (e.g., Google, news websites, blogs; 49.6% [642/1295]), and the GHS (48.7% [631/1295]). Participants were least likely to retrieve vaccine-related information from Twitter (13.7% [137/1295]), government officials (25.5% [330/1295]), Whatsapp (29.3% [379/1295]), and friends or family (33.4% [433/1295]).

By political affiliations (Table 4; Figure 2), participants consisted of NPP voters (elected; 42.3%), NDC voters (unelected; 12.6%), ‘Other’ voters (6.9%), and non-voters (26.7%). NDC voters (opposition) were more likely to be vaccine-hesitant than participants who did not vote (OR: 1.67; 95% CI: 1.07-2.59; *p =* .022). NPP voters were less likely to be vaccine-hesitant than participants who did not vote (OR: 0.57; 95% CI: 0.40-0.80; *p =* .001). A one-way ANOVA showed significant differences in ratings of trust in the COVID-19 vaccine between political groups (*F*(3,1143) = 16.69, *p* = .0001). Post hoc comparisons using the Tukey HSD test indicated that vaccine-related trust was significantly higher among NPP voters (*M* = 4.23, *SD* = 1.60) compared to NDC voters (*M* = 3.61, *SD* = 1.39), ‘other’ voters (*M* = 3.80, *SD* = 1.35), and non-voters (*M* = 3.71, *SD* = 1.35).

**Table 4.** Combined logistic regression model of factors contributing to COVID-19 vaccine hesitancy.

|  | OR | p-value | 95% CI |
| --- | --- | --- | --- |
| Age – older (40+) | 1.095 | .591 | 0.786 – 1.526 |
| Female | 1.594 | .008 | 1.130 – 2.250 |
| Urban community | 1.479 | .010 | 1.099 – 1.989 |
| Married or In a relationship | 0.886 | .458 | 0.644 – 1.219 |
| Higher education (undergrad or postgrad) | 1.649 | .002 | 1.210 – 2.248 |
| Being unemployed | 0.903 | .494 | 0.674 – 1.210 |
| Christian beliefs | 2.282 | <.000 | 1.486 – 3.506 |
| Political beliefs |  |  |  |
| National Democratic Congress (NDC, unelected) | 1.674 | .022 | 1.074 – 2.597 |
| New Patriotic Party (NPP, elected) | 0.575 | .001 | 0.408 – 0.809 |
| Other political party | 0.710 | .249 | 0.396 – 1.272 |
| Personally know somebody who received vaccine (Y) | 0.714 | .041 | 0.516 – 0.987 |
| Beliefs in misinformation | 1.274 | .089 | 0.864 – 1.683 |
| Uncertainty about misinformation beliefs | 1.865 | <.000 | 1.411 – 2.466 |
| Channels of COVID-19 information |  |  |  |
| Facebook | 0.907 | .546 | 0.660 – 1.246 |
| Whatsapp | 1.031 | .862 | 0.728 – 1.461 |
| Twitter | 0.956 | .842 | 0.617 – 1.484 |
| Mass media (e.g., radio, newspapers, TV) | 1.367 | .077 | 0.966 – 1.935 |
| Ghana Health Service or health workers | 0.692 | .020 | 0.508 – 0.943 |
| Government officials | 0.734 | .096 | 0.510 – 1.056 |
| Family members or friends | 0.864 | .383 | 0.623 – 1.200 |
| Internet (e.g., Google, news websites, blogs) | 1.388 | .032 | 1.029 – 1.872 |
| **Number of participants** |  |  | 1,138 |
| **R<sup>2</sup>** |  |  | 0.144 |

**Figure 2.**
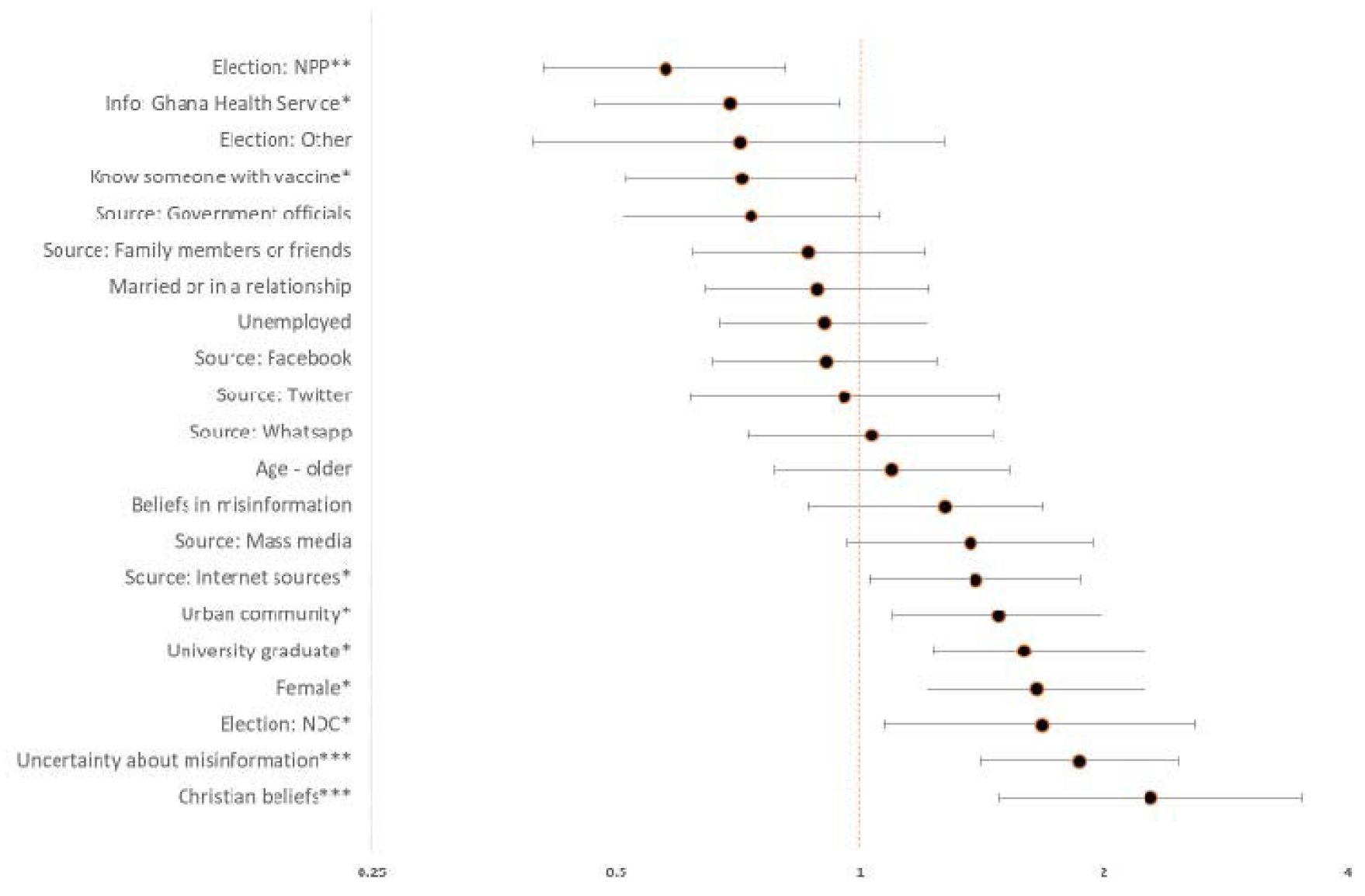
Summary of odds ratios and significance levels across factors studied (*\** < .05, ** < .01, *** < .001)

Participants who indicated agreement with at least one misinformation belief (i.e., participants who ticked ‘yes’ to indicate agreement) predicted marginally greater vaccine hesitancy compared to participants who did not indicate misinformation beliefs (OR: 1.27; 95% CI: 0.86-1.68; *p =* .089). However, participants who expressed uncertainty in at least one misinformation belief (i.e., those who ticked ‘I don’t know’ to indicate uncertainty) were significantly more likely to express vaccine hesitancy compared to participants who did not indicate uncertainty (OR: 1.86; 95% CI: 1.41-2.46; *p* < .000).

There were no significant predictors of vaccine hesitancy among participants who used social media platforms for COVID-19 vaccine-related information, such as Whatsapp (OR: 1.03; 95% CI: 0.72-1.46; *p =* .862) or Facebook (OR: 0.90; 95% CI: 0.66-1.24; *p =* .546) compared to participants who reported not using these platforms. However, participants who reported using Internet webpages (e.g., news websites, blogs) as a source of vaccine-related information were significantly more likely to report hesitancy than those who reported not using the internet (OR: 1.38; 95% CI: 1.02-1.87; *p =* .032). Participants who reported using the GHS as a source of vaccine-related information were less likely to report hesitancy compared to participants who did not report consulting the GHS (OR: 0.69; 95% CI: 0.50-0.94; *p =* .020).

There were several significant demographic and socio-demographic factors. Greater hesitancy was observed in Christian participants compared to Muslim participants (OR: 2.82; 95% CI: 1.48-3.50; *p =* .000); females compared to males (OR: 1.59; 95% CI: 1.13-2.25; *p =* .008); participants who completed more (vs. less) years of education (OR: 1.64; 95% CI: 1.20-2.24; *p =* .002); participants who lived in urban (vs. rural) communities (OR: 1.47; 95% CI: 1.09-1.98; *p =* .010); and participants who reported knowing somebody (vs. did not know somebody) who received the vaccine (OR: 0.71; 95% CI: 0.51-0.68; *p =* .041).

## Discussion

This study describes evidence of changes in overall levels of vaccine hesitancy in Ghana across four time-points during the COVID-19 pandemic. Hesitancy decreased between Survey 1 (August 2020) and Survey 2 (March 2021). This occurred around the time of vaccine availability and rollout, and amid extensive health promotion activity. However, hesitancy increased in Survey 3 (June 2021), which continued to increase further in Survey 4 (February 2022). Key reasons for refusing the vaccine included not having enough information about the vaccine and concerns about vaccine safety. Among key groups more likely to express hesitancy included Christians, urban residents, opposition party voters, females, and individuals who had completed higher education. It is possible that the increase in hesitancy rates observed in June 2021 may have been, in part, due to the global circulation of negative news stories surrounding the Oxford AZ vaccines at the time. The COVID-19 pandemic has been a global news story, and the actions of countries in the global north are seen and absorbed by those in the global south. Hesitancy rates during the pandemic elsewhere in sub-Saharan Africa vary greatly, ranging from 2.1% in Ethiopia, 17.3% in Malawi, and 35.5% in Mali (Kanyanda et al., 2021). However, our findings are comparable to hesitancy in many higher-income settings, including France (Detoc et al., 2020), South Africa (Lazarus et al., 2021), and USA (Fisher et al., 2020). A pragmatic viewpoint would be that vaccine hesitancy in sub-Saharan Africa, including Ghana, is neither higher nor lower than many other parts of the world.

A perceived lack of information and mistrust were common reasons for hesitancy, along with beliefs in pandemic-related conspiracy theories such as bioweapons, including mistrust in the Ghanaian government. Similar findings were also reported in Nigeria□ and Malaysia (Marzo et al., 2021). Voters of the unelected opposition party (NDC) in the Ghanaian general election were more likely to express hesitancy than voters of the elected political party (NPP). This was combined with lower confidence from NDC supporters in their level of trust in vaccine safety. Other studies have also demonstrated that political views impact views on vaccination. For example, vaccination rates were significantly lower in counties with a high percentage of US Republican voters (Albrecht, 2022), and French citizens who indicated voting for a far right candidate in the previous general election were more likely to state that they would refuse the COVID-19 vaccine if offered (Peretti-Watel, 2020).

Individuals who positively endorsed misinformation beliefs about COVID-19 contained weak effects on vaccine hesitancy. Thus, it is possible that anxiety caused by endorsing such imaginative beliefs are drivers of both vaccine acceptance and hesitancy. On the other hand, we found that individuals who indicated uncertainty with at least one misinformation belief about COVID-19 were more likely to express vaccine hesitancy. Perhaps uncertainty associated with one’s beliefs about the COVID-19 pandemic also translates across to beliefs about other man-made developments associated with the pandemic, including the vaccines (Bendau et al., 2021). Nevertheless, these findings should be interpreted with caution. Previous research has shown that the relationship between conspiracy theory beliefs and vaccine acceptance is highly complex, with various psychological dimensions mediating such beliefs and the propensity to get vaccinated, such as death anxiety (Simione et al., 2021).

Social media use was not a significant predictor of vaccine hesitancy despite WhatsApp and Facebook being two of the most widely used social media platforms in Ghana (Dukua-Sasu, 2020). However, the use of Internet webpages (e.g., news websites, blogs) was associated with hesitancy, suggesting that exposure to websites with potentially sensationalist stories about COVID-19 and vaccines – for example, news outlets that resort to sensationalism and exaggerated superlatives to remain competitive to advertising revenues – could influence individuals’ willingness to receive the vaccine (Ottwell et al., 2021). Further, using the GHS as a source of vaccine-related information was negatively associated with hesitancy, suggesting that information from official health sources may be key to building trust and countering misinformation.

In terms of religion, Christians were more hesitant than Muslims. A small number of churches have promoted anti-vaccine viewpoints, including the Christ Embassy, with headquarters in Nigeria and multiple churches in Ghana (Christ Embassy, 2021; Fani-Kayode, 2020). Previously in northern Nigeria, religious leaders developed misconceived perceptions about the polio vaccine. Many communities view religious leaders as a trustworthy and credible source of health advice and information, with research showing that religious leaders’ opinions can strongly influence social and behavioural norms (Adedini et al., 2018). However, for some religious individuals – particularly among those with strong beliefs of a controlling god – scientific inventions work against their core beliefs about the world (McPhretres & Zuckerman, 2018), and may highlight potential difficulties of persuading some religious leaders to promote pro-vaccination messaging to their followers. Given that the majority of participants in this study were Christians, it is not surprising that a large number of participants of Christian faith expressed doubts about vaccination. However, without in-depth examination, it is difficult to conclude whether Christianity is a true predictor in this study, or if it is the strength of conviction that they held about their religion.

Urban residents were more likely to be hesitant than rural residents, also reflected in research from Burkina Faso, Ethiopia, and Malawi (Kanyanda et al., 2021). Urban residents are typically more connected to the internet and social media and thus may be more exposed to vaccine-related misinformation than rural inhabitants who have fewer sources of information available to them. Thus, urban residents may be more worried about side effects and more likely to avoid vaccination, or are less afraid about COVID-19 due to having more access to information and daily alerts about COVID-19 (Ferdous et al., 2020; Islam et al., 2021; Kassim et al., 2021).

Finally, females were more likely to be hesitant than males, similar to findings from Nigeria (Jagede, 2007), and participants with more years of education were more likely to be hesitant than less educated participants, perhaps reflecting how more educated people in our sample are less likely to conform to social norms and behaviours (Adedini et al., 2018). This compares with surveys conducted in Burkina Faso, Ethiopia, Malawi, and Nigeria (Kanyanda et al. 2021). However, the opposite is true in research from Malaysia (Marzo et al., 2021) and Uganda (Echoru et al., 2021)

The core strength of this study relates to its relatively large number of participants, including temporal comparisons across four surveys. However, the need for internet access limits the representativeness of the sample population. Thus, certain demographic were under-represented, including individuals in rural areas and people of lower socio-economic status. Since our recruitment was also conducted using cross-sectional convenience sampling methods, there will also be presence of respondent bias. But the efficiency of data collection, the lower cost to advertise, and the acceptability of online survey recruitment may provide a useful alternative than formal regional or national surveys, especially during a global pandemic where new information from population surveys via remote or virtual methods may be urgently required. We also completed the same survey via in-person data collection in a rural area of the Oti region in January 2022 (Brackstone et al., 2022).

## 5. Conclusions

Hesitancy rates among unvaccinated individuals in Ghana continues to rise to worrying levels. However, vaccine awareness strategies are sensitive to subpopulation characteristics. Health promotion can use locally and nationally trusted knowledge providers (e.g., GHS) and disseminate good public health messaging via local trusted individuals – for instance, through religious groups – or on media platforms that are utilized by hesitant population groups. Many of these groups are reachable through targeted communication strategies, to which campaigns can focus on resolving concerns about vaccine-related side effects, and provide reassurance about the safety of approved COVID-19 vaccines to ensure high uptake and low vaccine hesitancy across Ghana.

## Data Availability

All data collected are available on the OSF

https://osf.io/r72hw/

## Author’s contributions

The study concept and design were developed by all the authors. KB coordinated the implementation of the study with support from all the authors. Analysis and interpretation of the data was conducted by KB, with input from KA, MJH, and LAB. Drafting of the manuscript was completed by KB with significant contributions from MH. Critical revision of the manuscript was performed by all the authors.

## Acknowledgements

We would like to thank Kingsley Osei and Kobby Nuamah (Opine World, Ghana), Hervé Akinocho (Center for Research and Opinion Polls, Togo), and Jean-Paul Fantognon (Togo Health Service) for their invaluable insight in this project throughout the past two years. We also thank the survey participants for their time and assistance in these surveys.

## Ethical approval

The surveys received ethical approvals from University of Southampton Ethics Committee (Institutional Review Board ID: 57267) and conformed to local ethical standards applied according to the Ghana Health Service (GHS), Ghana.

## Data availability

Data are available on the OSF: https://osf.io/r72hw/

## References

Adedini SA, Babalola S, Ibeawuchi C et al. Role of religious leaders in promoting contraceptive use in Nigeria: evidence from the Nigerian urban reproductive health Initiative. Glob Health Sci Pract. 2018;6(3):500–14.

Albrecht D. Vaccination, politics and COVID-19 impacts. BMC Public Health. 2022;22(96).

Bendau A, Plag J, Petzold MB et al. COVID-19 vaccine hesitancy and related fears and anxiety. Int Immunopharmacol. 2021.

Brackstone K, Attivor EJK, Boateng LA et al. Examining predictors of vaccine uptake and hesitancy in three rural sub-municipalities in Nkwanta South, Oti region, Ghana. 2022. doi: 10.6084/m9.figshare.19360988

Christ Embassy. Available from https://christembassy.org/pastor-chris-questions-the-impact-of-vaccines-on-recipients-in-five-years-time

Detoc M, Bruel S, Frappe P et al. Intention to participate in a COVID-19 vaccine clinical trial and to get vaccinated against COVID-19 in France during the pandemic. Vaccines. 2020;38(45):7002–06.

Dokua-Sasu, D. Most used social media platforms in Ghana as of the third quarter of 2020. Statista. 2020. Available from https://www.statista.com/statistics/1171534/leading-social-media-platforms-ghana/

Dotto C, Cubbon S. Disinformation exports: How foreign anti-vaccine narratives reached West African communities online. (2021). Available from: https://firstdraftnews.org/long-form-article/foreign-anti-vaccine-disinformation-reaches-west-africa/

Echoru I, Ajambo PD, Keirania E et al. Sociodemographic factors associated with acceptance of COVID-19 vaccine and clinical trials in Uganda: a cross-sectional study in western Uganda. BMC Public Health. 2021;21.

Fani-Kayode F. COVID-19 and the mark of the beast. Vanguard news Nigeria. 2020. Available from https://www.vanguardngr.com/2020/04/covid-19-and-the-mark-of-the-beast-femi-fani-kayode/

Ferdous MZ, Islam MS, Sikder MT et al. Knowledge, attitude, and practice regarding COVID-19 outbreak in Bangladesh: an online-based cross-sectional study. PLoS One. 2020;15.

Fisher KA, Bloomstone SJ, Walkder J et al. Attitudes toward a potential SARS-CoV-2 Vaccine. A survey of U.S. adults. Annals of Internal Medicine. 2020;173(12): 964–973.

Islam MS, Siddique AB, Akter R et al. Knowledge, attitudes and perceptions towards COVID-19 vaccinations: a cross-sectional community survey in Bangladesh. BMC Public Health. 2021;21.

Jegede AS. What led to the Nigerian boycott of the polio vaccination campaign? PLoS Med. 2007;4(3).

Kanyanda S, Markhof Y, Wollburg P et al. Acceptance of COVID-19 vaccines in sub-Saharan Africa: evidence from six national phone surveys. BMJ Open. 2021;11(12).

Kassim MAM, Pang NTP, Mohamed NH et al. Relationship between fear of COVID-19 psychopathology and sociodemographic variables in Malaysian population. Int J Mental Health Addict. 2021:1–8.

Lazarus JV, Ratzan SC, Palayew A et al. A global survey of potential acceptance of a COVID-19 vaccine. Nat Med. 2021;27(2):225–228.

Marti M, de Cola M, MacDonald NE et al. Assessments of global drivers of vaccine hesitancy in 2014: Looking beyond safety concerns. PLoS One. 2017;12(3).

Marzo RR, Ahmad A, Abid K et al. Factors influencing the acceptability of COVID-19 vaccination: A cross-sectional study from Malaysia, Vacunas. 2021.

McPhetres J, Zuckerman M. Religiosity predicts negative attitudes towards science and lower levels of science literacy. PLoS One. 2018;13.

Olapegba PO., Ayandele O, Kolawole SO et al. Preliminary Assessment of Novel Coronavirus (COVID-19) Knowledge and Perceptions in Nigeria. Social Sciences and Humanities Open. 2020.

Ottwell R, Puckett M, Rogers T et al. Sensational media reporting is common when describing COVID-19 therapies, detection methods, and vaccines. Journal of Investigative Medicine. 2021;69:1256–1257.

Peretti-Watel P. A future vaccination campaign against COVID-19 at risk of vaccine hesitancy and politicisation. The Lancet Infectious Diseases. 2020;20(7):769–70.

Puri N, & Coomes EA. Social media and vaccine hesitancy: new updates for the era of COVID-19 and globalized infectious diseases. Human Vac & Imm. 2020;16:2586–93.

Ritchie H, Mathieu E, Rodés-Guirao L et al. Coronavirus pandemic country profile. Published online at http://OurWorldinData.org. 2022. Available from https://ourworldindata.org/covid-vaccinations

Ritchie H, Mathieu E, Rodés-Guirao L et al. Ghana: Coronavirus pandemic country profile. Published online at http://OurWorldinData.org. 2022. Available from https://ourworldindata.org/coronavirus/country/ghana

Simione L, Vagni M, Gnagnarella C et al. Mistrust and beliefs in conspiracy theories differently mediate the effects of psychological factors on propensity for COVID-19 vaccine. Front. Psychol. 2021;12.

Wolter N, Jassat W, Walaza S. et al. Early assessment of the clinical severity of the SARS-CoV-2 omicron variant. MedRxiv. 2021.

